# Design and development of a flipped classroom-based online software course including online examination for medical statistics

**DOI:** 10.1101/2024.01.08.24300891

**Authors:** Rainer Muche, Andreas Allgöwer, Ulrike Braisch, Marianne Meule, Benjamin Mayer

## Abstract

Teaching medical statistics for students of human medicine should include both theoretical content as well as its implementation in practice using statistical software. The training period with a software may be time-consuming individually, so there is a risk that imparting methodological skills comes up short in such courses. The flipped classroom concept aims at students to familiarize themselves with both the statistical software and the theoretical content in an individual learning phase prior to the course. Then, within an attendance course deepening both aspects under the lecturer’s supervision is possible. Availability of the software as well as a variety of material to prepare the course are essential presumptions for the flipped classroom to be successful. We present the concept and implementation of an online flipped classroom course using the software SAS Studio. Students used their own devices (laptops or tablets) during the self-learning and examination phases, respectively. The pure online implementation in the flipped classroom setting enabled to contribute to an equalisation of the study plan. Moreover, even more students had the opportunity to participate in this practically oriented course in medical statistics. Initial evaluation results demonstrated that the course is well-accepted among students, while the students’ performance level was similar when compared to a non-flipped teaching approach.

## 1 Introduction

The teaching of biometric aspects in medical research, such as the planning of clinical and epidemiological studies and the application of medical statistics, should include not only theoretical content but also application-related components in the study of human medicine. The practical application of medical statistics is ideally taught with the help of statistical software. This allows students to demonstrate and try out for themselves how theoretical knowledge of statistical methods can be used to analyse data from practice. In accordance with our learning objectives, students should then be able to independently analyse the main features of research projects. Since the familiarization period in connection with the use of statistical software can sometimes be time-consuming for individuals, a traditional seminar structure with frontal teaching entails the problem that there may be less time available to explain theoretical knowledge, understanding of formulas and the interpretation of results in detail. As a result, individual follow-up work on the theoretical and practical content is sometimes necessary, but this learning phase is traditionally not accompanied by teachers. This poses a problem in that it is difficult for students to ask questions directly and any errors cannot be corrected by the teacher during the follow-up work.

### The Flipped Classroom-Approach

The teaching concept of the flipped classroom [5,6] reverses the typical teaching setting (learning in class and individual follow-up work afterwards): Students should familiarize themselves with the learning content before the lesson with the help of appropriate learning material. This also includes any exercises that may be part of the relevant course (e.g. in seminars). In the case of a statistical software course, this also concerns technical aspects of using the software (e.g. installation, uploading data, exporting results). In this way, students can initially prepare themselves according to their individual learning pace in the self-learning phase, so that during the subsequent face-to-face teaching phase, they can follow up and deepen their knowledge under the guidance of the teacher [5]. Due to a generally more homogeneous level of knowledge after self-learning, a software course in medical statistics also leaves than more time for the discussion of biometric aspects, such as the selection of suitable statistical methods and the interpretation of (study) results.

In general, the learning phases in the flipped classroom are divided into a self-learning phase outside of class and a subsequent attendance phase in class, which may be followed by a revision phase outside of class. For the self-learning phase, it is essential that suitable and varied learning material is available for students. Additional teaching elements intended for self-monitoring of learning progress, e.g. using multiple-choice questions or standard solutions to exercises, can also be offered. In a flipped classroom, the work phases of the face-to-face event must be structured differently than in a typical classroom setting. The following procedure is suggested in [5]:

1. address problems: What difficulties arose during the self-learning phase?
2. joint task processing: group work and discussions in the plenum.
3. active plenary: presentation of the results by the students.

In general, the successful implementation of a flipped classroom first requires that the students are technically able to access the learning material (e.g. via a learning platform). As the students have no direct contact with the lecturers in the initial self-learning phase, it must be ensured that appropriate support is available, e.g. in the form of consultation hours, forums or chat rooms. Teachers may therefore need to put a lot of effort into creating the learning material so that students have no problems grasping the content and lessons.

It has been shown that the flipped classroom is better accepted in compulsory courses than in elective courses [5]. The main advantages of the flipped classroom approach with regard to the course we present below can be summarized as follows [4,6]:

- students learn more intensively when they work independently
- both the learning pace and the learning strategy are determined individually by the students
- Students are usually more active
- interaction between students is encouraged through learning groups
- Attendance courses can focus on specific content (in this case biometrics and statistical software)
- teaching material can be used in the long term (during and after university)

### Aims of the article

In the following sections, we would like to present our concept for implementing the flipped classroom approach for a software course in medical statistics. We will present our conception and implementation of suitable teaching material as well as share our considerations regarding the selection of suitable statistical software. In particular, we will also present our concept of how an online exam can be integrated into such a course.

## 2 The concept of the flipped classroom software course in medical statistics at Ulm University

### 2.1 From face-to-face seminar to online flipped classroom

The basics of statistical analysis methods are taught in the interdisciplinary subject Q1/Medical Biometry, which is a compulsory course for all students in the curriculum in the human medicine program. At Ulm University, the course is part of the clinical section of this degree program and consists of a lecture (8 × 2h) and an accompanying seminar (6 × 2h), which was held with a maximum of 24 students per group before the introduction of the flipped classroom approach. In this seminar, students are required to attend and work on practice-oriented exercises using statistical software on the topics of experimental design, descriptive statistics, event time analysis, confidence intervals, regression, correlation and statistical hypothesis tests. We thus almost completely cover the learning objectives as formulated in the German National Competence-Based Learning Objectives Catalog for Medicine [16] for the field of medical biometrics. Prior to the introduction of the flipped classroom concept, each seminar was designed to begin with a brief review of the theoretical content of the respective topic and a presentation of the corresponding application options in the software used. This was followed by a software-supported, individual processing phase of exercises, the sample solutions of which were then presented and discussed in the plenary session. A graded short examination of 20 minutes concluded the seminar.

SAS Analyst [13], RExcel [15] and most recently SPSS have already been used in the context of this seminar concept. All of these programs used to date, despite their unrestricted availability in principle, showed various problems for comprehensive seminar preparation of the students. A corresponding recommendation for preparation was always made in order to achieve as homogeneous a level of knowledge as possible in classroom teaching, especially with regard to the practical handling of the evaluation program used, in the hope that this would leave sufficient time for the discussion of theoretical aspects. However, it often turned out that there was still too little time left for a recapitulation phase of what had been learned under the guidance of the teacher, so that it seemed sensible to switch the statistical software seminar to a flipped classroom approach. This has already been described for a biostatistics course [8], but had to be adapted to our seminar situation described above.

For the successful implementation of this flipped classroom seminar, which in particular also integrates our very well-evaluated concept of the examination based on six graded short examinations [9,14], various conditions had to be ensured, each of which is addressed in more detail below. Fundamentally, of course, this also included the willingness of all teaching staff involved to embrace the new teaching concept, as well as professional development in the area of teaching, which also had to cover the area of online teaching in particular. Appropriate further training was made possible by, among other things, attending courses at the Baden-Württemberg Center for Teaching and Learning and exchanging ideas with colleagues who have already successfully integrated the flipped classroom concept into their teaching [7].

The seminar, which is now offered as a flipped classroom, follows a slightly different structure compared to the previous face-to-face seminar. The entire seminar will be held online, especially the face-to-face sessions, which are currently held via Zoom. The main focus of the online presence is now mainly on presenting the necessary steps in SAS Studio to complete the exercises offered. Due to the integration of the flipped classroom concept, there is more time for the discussion of theoretical backgrounds during the online presence phase. The final short exam will also take place online using the online teaching platform described in section 2.3.

### 2.2 Selection of the statistical software

As part of an earlier research project dealing with the use of statistical software in applied teaching in medical biometry at a medical university, essential criteria for the selection of suitable software have already been defined, such as unrestricted use (duration of access license, costs), coverage of all relevant teaching content and comparatively easy access (preferably not programming-based) [12]. In the history of this statistical software course, we have already used various programs [12,13,15]. Our current choice fell on SAS Studio [1] due to the aspects briefly described below, although a different statistical program may be more appropriate at other locations due to the conditions and requirements there.

Depending on the existing infrastructure at our university (including room capacity in PC pools), the requirements already described above regarding the possibility of use (licenses and costs) and the availability of teaching content within the software as well as the experience with regard to the usage behaviour on the part of the students (almost without exception availability of laptops and tablets), the use of the SAS Studio software appeared to make the most sense in our setting. Here, cloud-based statistical analyses are possible on the basis of an intuitive user interface with the help of corresponding menus [1,18,19]. The user interface is divided into three parts and includes (i) a menu with the available data management and analysis functions, (ii) a menu that allows the assignment of the roles of the variables of interest for the intended analysis, and (iii) a pane where both the underlying SAS code (based on the settings of (ii)) and the results are displayed. By automatically generating the SAS code on which the analyses are based, this approach provides an additional entry point for students with an affinity for programming later on.

### 2.3 Teaching materials

The flipped classroom approach requires to make available learning materials in order to meet the requirements of autonomous preparation in the initial self-learning phase before the lesson. The aim should be to provide self-explanatory teaching materials in a variety of ways. Our intention was to provide the materials preferably in German (native language) in order to facilitate the use and access to the topics of biometrics and application of the software. Further open source video lectures (see section 2.3.2) on the use of SAS Studio were available in English.

Below we describe our approach to creating a wide range of teaching materials for our online software course in flipped classroom format.

#### 2.3.1 Moodle teaching platform

The Moodle learning platform is familiar to all students at our university and was also used to conduct our flipped classroom seminar [11]. The consistent use of Moodle was particularly strengthened by the COVID-19 pandemic, when most courses were held online. In addition to providing all the teaching materials we offer and conducting face-to-face courses online, we were also able to implement our online examination system (see section 2.5).

#### 2.3.2 Script

We believe that a coherent and summarizing textbook is important and helpful for students to familiarize themselves with statistical software. According to our search, only two English-language textbooks [2,3] were available regarding the use of SAS Studio in biostatistics. However, there was no German-language introduction to the use of SAS Studio for statistical analysis in the field of medical research, so we decided to publish a German-language textbook on the use of SAS Studio [1]. The creation of this textbook took about 18 months and the content covers all relevant steps of both data management and analysis with the menus of SAS Studio that are relevant for our course in medical statistics. All needed applications are explained in text-based form and supported by corresponding screenshots, so that users also have the opportunity to visually understand applications directly and compare them on their own screen.

#### 2.3.3 Educational videos

Students on our course can also access short instructional videos that introduce the use of SAS Studio in relation to various methodological topics. Some of these videos were already available in English and could be accessed directly via SAS [19]. In addition, we produced similar videos in German, some of which also addressed other methodological topics, such as creating bar charts or performing a Kaplan-Meier survival analysis.

All videos were recorded with the software OBS Studio (Open Broadcast Software Studio [17]) and cut and edited with the free video editor software Shotcut [20] by two members of our working group. The familiarization and use of these two software products is intuitive and does not require a great deal of time. Before each recording, an outline of the content of the video to be produced was created. From a didactic point of view, only short videos of a maximum length of 5 minutes were created in order to keep the students’ concentration high. Each video therefore only contains one specific aspect of the use of SAS Studio. This resulted in a total of 25 videos (14 in German and 11 SAS videos), which are now available on our Moodle page and thematically reflect the teaching content of our seminar.

#### 2.3.4 Exercises and self-tests

Practical exercises have been developed for each of the methodological topics covered in the online classroom course, which are to be completed in the preceding self-learning phase using the SAS Studio software (Figure 2a). For this the students get real data from a clinical project [21]. To enable self-directed learning, students can access the solutions, which are available a few days after the exercises have been published. These exercises are important for the examinations, as the type and level of difficulty of the examination questions are based on them.

To further expand the opportunities for self-directed learning, we have also implemented multiple choice-based self-tests for each topic block. These can be integrated using the Moodle activity “Test”.

### 2.4 Online implementation of the seminar

As described, students should work independently on their tasks and the use of SAS Studio in accordance with the flipped classroom approach before each attendance phase. The self-developed solutions and any questions, which may concern both the use of the statistical software and the theoretical background of the respective topic block, are then discussed together with a teacher during an online attendance session. Around 30-40 students are required to attend. A Zoom link is made available on Moodle for the course in online presence, which can only be used by students enrolled in a particular course.

At the beginning of each online presence event, students are given the opportunity to ask questions about any problems they may have encountered when using the statistics software or working through the exercises in general. The results obtained are then discussed in the plenary session using sample solutions. Further possible solutions can be presented and discussed by the students. In this context, a strong focus is placed on the interpretation of the content of the exercises. After a further opportunity to ask any remaining questions, each of the six seminars is then concluded with a short online examination (see section 3.4).

### 2.5 From the previous standard to the online exam

For some years now, our examination concept for the medical statistics seminar described here has not included a written examination at the end of the semester. Instead, students write a short examination lasting 20 minutes at the end of each seminar. The cumulative total score of all short exams is then decisive for the grade awarded; the pass mark is currently 60% of the total score. We have opted for this form of examination during the semester because the content of our seminar builds on each other and in this way continuous learning can be achieved and, as a result, learning success can be improved. As part of the short examination, students work on smaller tasks, if necessary using the statistical software they have learned, whose content and level of difficulty are based on the previously discussed exercises. For this purpose, real data from a clinical sample data set is used, which differs individually for each student by randomly drawing a partial data set. Details on the specific implementation of the examination format with subsequent semi-automatic correction were described in [14]. The short examinations are held as so-called open book examinations, i.e. students are allowed to use their course materials to complete the tasks. Due to the very tight time limit, students are forced to be well prepared despite the open book principle in order to complete the exam within the given time frame. As this examination concept was very well accepted and evaluated by students [14], our aim was to retain this examination concept in the flipped classroom online format described here.

#### 2.5.1 Online exam in Moodle

The Moodle learning platform is used for our online exam concept, as well as for the online face-to-face event. As the short exam forms the conclusion of each online seminar, students can remain in the Zoom session that is already in progress. The “Test” application is used in Moodle for the short exam, which is described in more detail below. The examination records to be processed individually have an ID and are provided via Moodle in Excel format. At the start of the semester, each student is assigned an ID that determines which data record must be processed. In concrete terms, the short examination procedure is as follows: The students

- switch on their camera in the current zoom session
- make sure that SAS Studio is open
- download their individual data set to be used from Moodle
- import the now locally available Excel data set into SAS Studio
- start the test application in Moodle, in which they can enter the results of their analyses and other answers within a maximum of 30 minutes

Text fields can be used to draw attention to the most important regulations for the test and instructions can be given on how to conduct the test. Students can also be asked for their formal consent to the test in this way before the test application actually starts the short test. When the test starts, automatic time recording begins for each student individually; after this time has elapsed, the test application can no longer be edited automatically.

The “Test” application in Moodle makes it possible to implement questions and corresponding solution fields for answering them. The input options can be restricted so that, for example, only certain values are accepted. The “Test” application allows four different input field modes to be used (multiple choice (MC), numerical input fields, text input fields (e.g. for interpreting results) and uploads, e.g. to provide created graphics as a solution file). In the case of MC questions, students can select the answer directly; in the case of results generated with SAS Studio, the results must be entered manually. Although the usual copy-paste can be used for this, it should be noted that Moodle only allows the comma as a decimal separator, whereas in SAS Studio the point is output as a decimal separator. If this is overlooked by the students, a corresponding note will automatically follow before the official completion of the “Test” application by Moodle itself. Navigation through the test is made easier for students by a corresponding overview, which also shows the time still available. There are special markings for questions that are currently being processed, that have already been completed or that still need to be processed.

All entries within the launched “Test” application are saved by Moodle in a database and can be exported in various formats for further processing.

After several prototype test runs within our working group, the online examination system described here was implemented for the first time in winter semester 2022/23. A total of 99 students were registered for the seminar described here and subsequently assessed the course (see section 3).

#### 2.5.2 Organizational and legal aspects

Completing the online exam with the camera switched on in the Zoom session should help to ensure that the short exam is actually carried out independently and only with the help of the permitted aids. Although this approach can also only be used to carry out an independent examination to a limited extent, verbal communication between students is at least prevented.

In the event of technical problems or questions about the content of the short exam, students can communicate with the teacher via the chat function in Zoom. However, the chat room settings stipulate that students cannot communicate with each other, only with the teacher.

Automated registration for the seminar and the data protection-compliant publication of the points achieved and the grades obtained have also been developed for some time [14] and can be used in the online flipped classroom described above. Each student receives an email with a numerical code at the start of the semester, which acts as a key for publishing points and grades on Moodle. This relatively simple and quick way of publishing points makes sense for didactic reasons, as we want to inform students of their accumulated points as early as possible before the next seminar date for the semester-accompanying examination via 6 short tests.

The use of the examination form described here must be formally and legally secure. To this end, our examination form of short examinations during the semester was agreed with the Dean of Medicine at our university. The exemptions for examinations in the course of the coronavirus pandemic also made it possible to convert the examination concept to the online version described. Following the pandemic, the currently valid study and examination regulations of our university for the Human Medicine degree program (preclinical and clinical study section) stipulate that examinations can also be taken in electronic form in principle. The general study and examination regulations for Bachelor’s and Master’s degree programs at Ulm University dated 13th July 2022 also provide for online examinations, which essentially correspond to our approach.

## 3 Evaluation

Before we brought our developed flipped classroom concept into regular use as part of the statistical software course described here, the course was first offered as an elective for students of human medicine at Ulm University in the winter semester 2020/21. Our aim here was to evaluate the feasibility and student acceptance of the course material described above. A total of 26 students took part in this course, 15 of whom took part in the evaluation. Their results were presented in detail in a separate publication [10]. Even taking into account the very small sample size, the feedback was generally very positive, with the majority (14 out of 15) considering the course in the format offered to be an appropriate alternative to the regular course format described in section 2.1.

### 3.1 Evaluation and learning success during the first regular use of the online flipped classroom

In the winter semester 2022/23, the Q1/Biometrics seminar was offered to 247 students in the 7th semester of the Human Medicine degree program, 99 of whom attended the seminar using the online flipped classroom presented. For these 99 students, a 90-minute introductory event was offered before the official start of the seminar, during which the didactic idea of the flipped classroom format was presented and an introduction to the SAS Studio statistical software was given.

The overall grade in the official faculty evaluation of all courses was very good for the Q1/Biometrics seminar with an average score of 5.4 (SD=0.7), whereby this evaluation is based on a six-point scale. However, this evaluation result relates to the total number of n=216 students who submitted an evaluation for the Q1/Biometrics seminar. A separate analysis of the regular seminar and flipped classroom groups is unfortunately not planned or possible on the basis of this evaluation, meaning that specific evaluation results for the online flipped classroom course are not available.

However, our own data on the total number of points achieved by the students and, consequently, their grade distribution show that there are no differences in terms of grade distribution between the regular seminar groups and the flipped classroom approach. We can therefore at least assume that the method of implementation and online examination has no influence on the grade in Q1/Biometrics.

In addition, some excerpts from the evaluation of the PC course can be reproduced here:

Positive aspects:

- Really sensible/good idea to offer the PC course; it teaches you how to use a statistics program, which you might really need at some point (e.g. for promotion)
- I also think the idea of the short tests is very good, as you can always learn and understand a small part and don’t have to write another exam at the end of the semester in addition to the 10 others
- I didn’t find it bad at all that the PC course was held online. Everyone has to sit at their laptop anyway. You could keep it that way in the future. Personally, I also think it’s better than doing it in the PC pool.

Negative aspects / room for improvement:

- Operating 3 programs at the same time and hoping the Internet connection remains stable is unnecessary stress.
- At the beginning I was a bit disappointed because I had to teach myself everything anyway due to the flipped classroom principle and the novelty of using SAS Studio - which I could do without taking the course. I would have liked a bit more of an introduction to the program from the lecturers, especially at the beginning.

When reviewing the open questions, it is noticeable that there are no direct critical comments on the online examination situation. Detailed questions about the difficulty, grading or timing of the exams are mentioned, but no general comments. Overall, we can be satisfied with the implementation of the course after this evaluation, also because no problems or objections have been raised by students during the implementation of the online examinations to date.

## 4 Discussion and outlook

This article presents the design and implementation of a course for learning basic biometric skills and the use of statistical software as part of this course using the flipped classroom approach. This compulsory course in the cross-sectional subject Q1/Biometrics has since been offered in the human medicine program at Ulm University. In order to maximize the learning success and the understanding of the importance of the subject, online presence events with sufficient time for discussion and interpretation of the analysis results are very important. Therefore, the self-study phase outside of the lecture provided for in the flipped classroom approach will improve students’ prior knowledge before the face-to-face seminar, leaving more time for in-depth discussion of biometric content. We have seen that such a concept works in an online setting and is accepted by students before our first implementation in the context of regular compulsory teaching based on a corresponding elective subject.

The prerequisites for the use of the flipped classroom concept are provided by the use of suitable statistical software and various learning materials. SAS Studio offers unlimited and free access to all relevant statistical methods that are formulated in accordance with the learning objectives of the National Competence-Based Learning Objectives Catalog for Medicine. A German textbook has been published and serves as an introduction to the practical application of the software for our students. The availability of further teaching material (exercises, teaching videos, self-tests) was evaluated as very helpful, and the Moodle learning platform also made it possible to implement an online examination.

The advantage of students attending the seminar using their own devices is that they can get used to the individual circumstances (e.g. operating system-related differences in the use of the SAS Studio interface) and are already familiar with the software environment in the event of any future projects. During the development process, we considered offering advice to students who have individual problems using the software or setting up access. However, it turned out that there was no need for this. Isolated problems could be solved directly during the attendance phases.

The evaluation results presented are not yet available in the level of detail that would be necessary for a comprehensive assessment of the online flipped classroom presented, especially with regard to a comparison with student groups who do not attend the compulsory Q1/biometrics seminar using statistical software. This should ideally be carried out on the basis of a cluster-randomized study, in which the achievement of learning objectives should be specifically recorded.

We have presented our concept and the implementation of an online flipped classroom for a statistics software course for the introduction to medical statistics. We hope that this contribution will also provide an incentive to reflect on didactics in the context of teaching in our subject. Based on the initial feedback, we are optimistic that the approach presented can help both students and teachers to benefit from it in the long run.

## Data Availability

All data produced in the present work are contained in the manuscript

## Conflicts of interest and financing

The authors declare that they have no conflicts of interest in connection with this article.

This work was supported as a teaching project by the Medical Faculty of Ulm University under the funding code L.LPB.0001.12_2022.

